# Occupational and Environmental Challenges and Effects of COVID-19 Testing Implementation Experienced by HIV Viral Load Laboratory Staff within a Public Health Sector Laboratory in South Africa

**DOI:** 10.64898/2026.02.16.26346134

**Authors:** Somayya Sarang, Eunice Matingo-Mutava, Naseem Cassim

**Affiliations:** National Priority Programmes (NPP), National Health Laboratory Service (NHLS, Johannesburg, South Africa; Faculty of Health Sciences, University of Johannesburg, Johannesburg, South Africa; Wits Diagnostic Innovation Hub (Wits DIH), Faculty of Health Sciences, University of Witwatersrand, Johannesburg, South Africa

**Author notes:** **Corresponding author:** Naseem Cassim, Sunnyside Office Park, Building B, 1^st^ Floor, 32 Princess of Wales Terrace, Parktown, Johannesburg, 2193.

**Keywords:** Burnout, COVID-19 Testing Implementation, Occupational and Environmental Challenges, Psychological Injury, staff workload, HIV Viral Load Laboratory Staff, Workforce Retention

## Abstract

**Background:** The COVID-19 pandemic required South African public sector HIV viral load (VL) laboratories to scale up Severe Acute Respiratory Syndrome Coronavirus 2 (SARS-CoV-2) testing while maintaining essential HIV services. This placed additional pressure on diagnostic services. This dual mandate introduced significant occupational and environmental challenges (OEC) for staff that remain underexplored.

**Objective:** This study aimed to investigate the OEC and effects that staff experienced during the implementation of COVID-19 testing at public sector VL laboratories in South Africa.

**Methods:** A quantitative, cross-sectional study utilised a census approach among technical and support staff. Data were collected via a structured REDCap questionnaire using 5-point Likert scales. Pre- and post-implementation challenges were assessed across four domains: workload, environmental conditions (space, ventilation, waste), communication, and PPE availability. Statistical analyses included the Wilcoxon Signed-Rank and Spearman’s correlation tests.

**Results:** Perceived occupational challenges increased significantly across all domains post-implementation. Staff workload saw the highest rise (mean score 3.02 to 3.53). Adverse health effects were pervasive; 80.2% of staff reported burnout/fatigue, and 76.5% reported increased anxiety/stress. A strong positive correlation was observed between post-COVID-19 challenges and adverse mental and physical health outcomes (rho = 0.449, p < 0.001). Furthermore, 35.8% of staff considered resigning due to increased job demands.

**Conclusion:** Integrating COVID-19 testing exacerbated systemic weaknesses, causing measurable psychological injury and threatening workforce retention. Findings suggest that the diagnostic workforce requires formal crisis surge staffing models and institutionalised mental health support to safeguard personnel and maintain essential services during future health emergencies.

## Introduction

Globally, the public-sector healthcare sector, particularly diagnostic laboratory staff, faced significant challenges during the Coronavirus 2019 (COVID-19) pandemic. Diagnostic laboratories are crucial for timely diagnoses to mitigate the spread of viruses.^[1]^ South Africa has an extensive Human Immunodeficiency Virus (HIV) epidemic, with an estimated eight million people living with HIV (PLHIV).^[2]^ HIV viral load (VL) testing is crucial in this context, monitoring the effectiveness of antiretroviral therapy (ART) and ensuring viral suppression, which is essential for meeting global Joint United Nations Programme on HIV/AIDS (UNAIDS) 95-95-95 targets.^[3, 4]^ The public sector National Health Laboratory Service (NHLS) conducts over six million VL tests annually through a network of centralised laboratories.^[5]^ During the pandemic, these VL laboratories operated under a challenging dual mandate: expanding capabilities to conduct Severe Acute Respiratory Syndrome Coronavirus 2 (SARS-CoV-2) testing while continuing to offer essential HIV monitoring services.^[6]^

This integration of mass COVID-19 testing protocols introduced specific operational obstacles, amplifying existing occupational and environmental pressures. While the vital role of these laboratories is acknowledged^[6]^, their specific experiences remain underexplored, particularly regarding the occupational and environmental challenges (OEC) and their impact on mental and physical health. Current research often focuses broadly on public health responses, rather than the granular operational context within diagnostic laboratories. There is even less research specifically looking at the impact on high volume molecular laboratories offering both VL and COVID-19 testing. It remains unclear in this setting what the OEC impact was specifically in VL laboratories with high testing volumes adopting a centralised model. It would be critical to understand these OEC in the context of pandemic preparedness and maintaining and sustaining HIV diagnostic services.

The investigation was conceptually guided by the Health Belief Model (HBM), which frames OECs as ‘perceived barriers’ preventing staff from maintaining a healthy work-life, while the resulting health effects are viewed as ‘Perceived Severity/Susceptibility’.^[7]^ The global diagnostic sector reported widespread OECs stemming from the pandemic response. The most pervasive challenge was the overwhelming workload, leading to relentless work hours and extended shifts.^[8]^ This systemic strain acts as a potent perceived barrier within the HBM. Resource scarcity was acute; global reports indicated widespread personal protective equipment (PPE) shortages, which generated widespread anxiety among workers by increasing their perceived susceptibility to infection.^[9–11]^ Furthermore, issues like maintaining adequate ventilation, managing increased infectious waste, and securing laboratory space added environmental pressure.^[10, 12]^ Organisational support, including clear communication regarding protocols, was often inconsistent, acting as a barrier to self-efficacy.^[13, 14]^

The accumulated systemic strain translated directly into adverse psychological and physical health outcomes.^[8, 15]^ Studies identified high rates of burnout, anxiety, and depression among healthcare workers.^[8, 15]^ For laboratory staff, this stress was exacerbated by the fear of perpetuating infection through sample handling, combined with feeling invisible compared to frontline clinical staff.^[16]^ Chronic stress and inadequate support reduce job satisfaction, driving staff to consider leaving the profession.^[15]^ Physical symptoms such as chronic headaches, musculoskeletal pain, and fatigue were also associated with prolonged use of cumbersome PPE.^[17, 18]^

South Africa’s context is unique due to the immense scale of the National Health Laboratory Service (NHLS) with a dual mandate to sustain VL monitoring for millions of PLHIV while urgently scaling up Severe acute respiratory syndrome coronavirus 2 (SARS-CoV-2) testing.^[2, 6]^. The NHLS provides diagnostic services for over 80% of the population in South Africa.^[5]^ Despite the extensive molecular capacity and experience that has been built up from 2004 to manage high VL volumes from a large HIV program, significant challenges were encountered during its rapid transition to large scale SARS-CoV-2 testing.^[5]^ During the pandemic, VL volumes ranged from 5,706,02 (202) to 5,788,673 (2021).^[19]^ A local study reported that during waves one and two, up to 40% of the VL volumes at the time were for SARS-CoV-2 testing, emphasising the significant increase in workload.^[6]^ The World Health Organization (WHO) has reported that there were 4,073,151 COVID-19 cases in South Africa, the majority of whom were diagnosed within the NHLS. This operational burden, amplified in a resource-limited public health setting, warrants focused investigation.^[20, 21]^

The purpose of this study was to investigate the specific OEC’s and subsequent adverse health outcomes experienced by public-sector VL laboratory staff following the implementation of COVID-19 testing in South Africa. The study will provide evidence for OEC, given the potential impact on diminished mental health, increased error rates, and compromised patient care. This study aimed to investigate the OEC faced by laboratory staff pre- and post-implementation of COVID-19 and to detail the consequences for staff’s health and well-being. Furthermore, this research aims to assess the impact of this new testing workload on mental and physical health, particularly in relation to stress and job satisfaction stemming from increased workloads and safety concerns.

## Methods

### Ethical considerations

The study strictly adhered to all ethical guidelines and protocols prescribed by the University of Johannesburg (UJ) and the National Health Research Ethics Council (NHREC) of South Africa. Approval was received from the UJ Higher Degrees Committee prior to data collection. Formal ethical approval, with clearance number: REC-3556-2025, was secured from the UJ Research Ethics Committee. Respondents provided electronic informed consent via a tick box on REDCap before completing the questionnaire. Respondents who declined consent were unable to proceed with the survey, achieved by using branching logic. The survey was completely anonymous, with identifying details, such as names, employee numbers, or Internet Protocol (IP) addresses, not requested (S1). The data handling adhered strictly to the Protection of Personal Information Act (POPIA). The research was self-funded, and the authors declared they received no financial support for the research, authorship, and/or publication of this article. Furthermore, the authors declared no financial or personal relationship(s) that may have inappropriately influenced them in writing the article.

### Study Design

This study employed a quantitative, causal-comparative/quasi-experimental and cross-sectional study design. This design allowed for the comparison of conditions across two distinct time points: pre- and post-COVID-19 testing integration.

### Study Setting

The study was conducted across 17 centralized Viral Load (VL) laboratories within the South African National Health Laboratory Service (NHLS) diagnostic network, spanning eight provinces: Gauteng, Limpopo, Mpumalanga, Eastern Cape, KwaZulu-Natal, Western Cape, North West, and the Free State. This network, which provides approximately 80% of South Africa’s pathology services, proved pivotal in the rapid scale-up of COVID-19 molecular testing.^[5, 6]^ These specific facilities were selected as they represent a resource-limited, high-volume molecular diagnostic environment that managed a “dual mandate” crisis: maintaining essential HIV monitoring while absorbing the massive capacity demands of the pandemic response. The study population comprised 100 technical and support staff involved in COVID-19 testing during the acute phase of the pandemic (2020–2021), a period characterized by four distinct epidemiological waves. To facilitate this surge, the laboratories leveraged existing high- and medium-throughput automated molecular platforms, including 14 Cobas 8800 (Roche Diagnostics, Basel, Switzerland) and 30 Alinity m (Abbott Diagnostics, Chicago, IL, USA) systems. These platforms, originally placed via national reagent agreements for HIV VL monitoring, were strategically repurposed to integrate qualitative SARS-CoV-2 assays, utilizing their established scale to meet emergency diagnostic needs

### Study population and sampling

A census sampling approach was utilised, inviting all eligible staff to participate. The population included all eligible technical and support staff, comprising laboratory assistants, medical technicians, medical technologists, medical scientists, pathologists, and laboratory managers employed at the 17 centralised VL testing laboratories, who were involved in or directly affected by the implementation of COVID-19 testing. The target population included staff who worked during the primary phase of the pandemic response (2020–2022), when OECs were most acute, and was estimated to be approximately 100 laboratory staff. The minimum required sample size was calculated as 79, assuming a confidence interval of 95% and a margin of error of 5%.

### Data Collection and Analysis

The invitation to participate in the study was sent via email on the 4^th^ of July 2025. Responses were received between the 15^th^ of July and the 20^th^ of August 2025.

Data were collected via an electronic self-administered questionnaire (SAQ) hosted on the secure REDCap platform (S1).^[22, 23]^ The questionnaire employed 5-point Likert scales to measure OECs pre- and post-COVID-19 implementation, encompassing domains like staff workload, PPE availability, environmental conditions, and communication/support. The questionnaire was broken up into four sections as follows: (i) A: demographic characteristics which included gender, age category, level of education, registration with a professional board and years of registration, (ii) B: OECs pre-COVID-19 testing that included challenges with equipment availability, staff workload, laboratory safety protocols, PPE, environmental conditions, communication and support from laboratory management, psychological factors and training, (iii) C: OEC’s post-COVID-19 testing and (iv) D: Mental and Physical Effects of Implementation (MPEI) that assessed stress/anxiety levels, burnout, fatigue experienced, physical and mental health affected, job satisfaction and consideration about leaving their jobs (S1).

### Methodological Rigour and Bias Mitigation

To ensure data integrity and mitigate measurement bias, the research instrument underwent a rigorous validation process. Content and face validity were established by aligning questionnaire items with Health Belief Model (HBM) constructs and established literature [^[17, 24]^, followed by a formal review by subject matter experts to ensure technical accuracy and relevance to the HIV viral load laboratory context. Criterion validity was ensured by mapping items against the study’s specific objectives to prevent construct under-representation. Instrument reliability was confirmed through internal consistency testing; all multi-item scales, including workload and burnout, met the predetermined Cronbach’s alpha threshold of 0.70.

### Statistical analysis

Exploratory Factor Analysis (EFA) was performed to confirm SAQ construct validity. Statistical computations were performed using the Statistical Package for the Social Sciences (SPSS), version 30 (IBM, Armonk, NY, USA).^[25]^ Descriptive statistics were used to profile the respondent profile as well as the extent of the challenges. Frequencies and percentages were generated for all socio-demographic factors (confounders), including gender, age, education level, professional registration and years of experience. For Likert scales coded from 1 to 5, means (M) and standard deviation (SD) were reported, including the composite scores for each OEC domain and outcome variable. Post-COVID-19 challenges were merged into a single, overarching source of pressure, namely the Unified Crisis Strain (AIC). For the quantitative mean scores (M) derived from the 5-point Likert scale for both individual items and the composite scores (OEC/AIC and MPEI), a qualitative classification schema anchored at the neutral midpoint of 3.0 was employed. Specifically, mean scores ranging from 3.41 to 5.00 were interpreted as indicating a High/Severe Challenge or Negative Effect, reflecting strong collective agreement among respondents.^[26]^ Mean scores ranging from 2.61 to 3.40 indicated a moderate or significant challenge, particularly when scores confirmed the presence of a negative outcome, such as the Composite Health Effects Score (MPEI), which significantly exceeded the neutral threshold. Finally, mean scores below 2.60 were generally classified as a ‘Low or Negligible challenge’.^[26]^ Given that most variables were not normally distributed (*p* < 0.05), non-parametric statistics, including the Wilcoxon Signed-Rank test to compare pre- and post-implementation challenge scores. Spearman’s rho (ρ) non-parametric correlation analysis was employed to robustly examine the relationship between the composite AIC and the MPEI.

## Results

### Instrument Reliability and Validity (EFA)

The study successfully recruited 81 respondents (N=81), achieving an 81% response rate. The structural integrity and construct validity of the research instrument were established through Exploratory Factor Analysis (EFA) and internal consistency testing. Sampling adequacy was confirmed by Kaiser-Meyer-Olkin (KMO) values of 0.782 for the pre-COVID-19 period and 0.887 for the post-COVID-19 period, supported by a significant Bartlett’s Test of Sphericity (p < 0.001). In the pre-pandemic analysis, two distinct factors emerged: Infrastructure/Biosafety (OEC F1) and Workforce/Support (OEC F2), explaining 58.9% of the total variance. However, post-pandemic data revealed a critical structural shift toward a single-factor model, termed Unified Crisis Strain (AIC), which accounted for 62.4% of the variance; this transition suggests that the pandemic effectively fused previously discrete operational challenges into one overarching systemic pressure. Furthermore, the outcome measure, Mental and Physical Effects of Implementation (MPEI), loaded onto a single factor identified as Psychological Injury. The instrument’s reliability was evidenced by a robust overall OEC scale (Cronbach’s Alpha = 0.84), with all multi-item sub-scales consistently exceeding the 0.70 threshold to provide a rigorous foundation for subsequent inferential analysis.

### Demographic Profile and Instrument Reliability

The workforce was predominantly female (n=57,70.4%), mostly aged 30 to 49 years (72.8%), and the largest professional category was registered medical technologists (70.4%) and technicians (11.1%) (Table 1). The majority of staff obtained a Bachelor of Technology (38.3%), followed by a postgraduate degree (24.7%). Crucially, 71.6% of the sample possessed significant experience (5 to 19 years)

**Table 1:** Demographic characteristics of respondents for a study that assessed Occupational and Environmental Challenges and Effects of Coronavirus Disease 2019 (COVID-19) testing implementation experienced by HIV viral load laboratory staff within a public health sector Laboratories in South Africa.

| Category | N= (%) |
| --- | --- |
| Overall | 81 (100.0) |
| **Gender** |  |
| Female | 57 (70.4) |
| Male | 21 (25.9) |
| Prefer not to state | 3 (3.7) |
| **Age Category** |  |
| 20–29 | 6 (7.4) |
| 30–39 | 36 (44.4) |
| 40–49 | 23 (28.4) |
| 50–59 | 12 (14.8) |
| 60+ | 4 (4.9) |
| **Highest Education** |  |
| Matric certificate | 6 (7.4) |
| National Diploma | 18 (22.2) |
| Bachelor of Technology | 31 (38.3) |
| Undergraduate Degree | 6 (7.4) |
| Postgraduate Degree | 20 (24.7) |
| **HPCSA Registration Category** |  |
| Laboratory Assistant | 1 (1.2) |
| Medical Technician | 9 (11.1) |
| Medical Technologist | 57 (70.4) |
| Medical Scientist | 4 (4.9) |
| Pathologist | 7 (8.6) |
| Other | 3 (3.7) |
| **Years Registered with HPCSA** |  |
| <5 years | 4 (4.9) |
| 5–9 years | 20 (24.7) |
| 10–19 years | 38 (46.9) |
| 20–30 years | 12 (14.8) |

HPCSA: Health Professions Council of South Africa COVID-19: Coronavirus Disease 2019

### OEC analysis

For staff workload, the high pre-COVID-19 mean of 3.02±0.71 (indicating a mild challenge) increased to 3.53±1.16 post-COVID-19 (Table 2).

**Table 2:** Analysis of the mean pre- and post-Coronavirus Disease 2019 (COVID-19 scores) assessed for occupational and environmental challenges experienced by HIV viral load staff within public health sector laboratories in South Africa. Data was assessed using a likert scale (1 to 5), with the mean and standard deviation (SD) reported.

| Variable | Pre-COVID-19 Mean (SD) | Post-COVID-19 Mean (SD) |
| --- | --- | --- |
| Equipment availability | 2.52 (0.78) | 2.59 (0.79) |
| Staff workload | **3.02 (0.71)** | **3.53 (1.16)** |
| Laboratory safety protocols | 2.41 (0.83) | 2.63 (0.98) |
| PPE availability | 2.49 (0.83) | 2.72 (1.04) |
| Environmental conditions (space, ventilation, waste) | 2.48 (0.83) | 2.81 (1.10) |
| Communication/support from management | 2.54 (0.83) | 2.81 (1.16) |

PPE: Personal Protective Equipment COVID-19: Coronavirus Disease 2019 SD: Standard Deviation

Workload was the most frequently reported challenge, with 64.2% of respondents agreeing or strongly agreeing that it was a problem (Figure 1). Concerns regarding PPE availability were reported by 51.9% of staff, with the mean increasing from 2.49±0.83 (pre-) to 2.72±1.04 (post-COVID-19). For the other variables, the post-COVID-19 mean ranged from 2.59±0.79 (equipment availability) to 2.81±1.16 (communication/support from management). A Wilcoxon Signed-Rank Test: confirmed that the increase in challenge scores from pre- to post-implementation was statistically significant across domains (p < 0.05).

**Figure 1:**
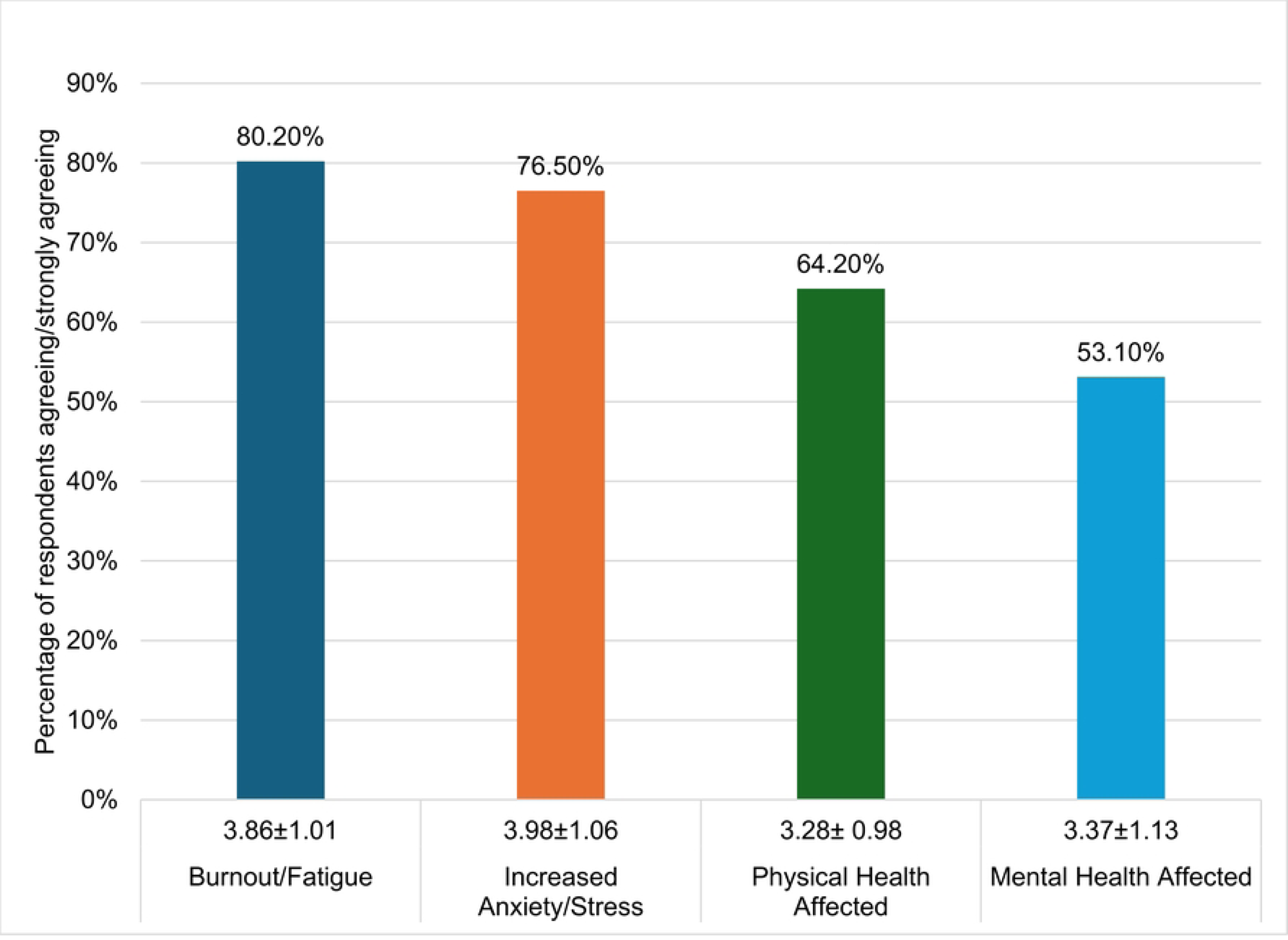
Analysis of the percentage of staff who agreed or strongly agreed with the Mental and Physical Health Effects of Implementation (MPEI) effects of Coronavirus 2019 (COVID-19) implementation relating to burnout/fatigue, anxiety/stress, and physical and mental health. This is based on responses by staff at HIV viral load laboratories in the public health sector, South Africa. COVID-19 Coronavirus Disease 2019 MPEI: Mental and Physical Health Effects of Implementation

### MPEI analysis

As shown in Table 3 and Figure 1, the majority of respondents reported experiencing burnout/fatigue (80.2%) (M=3.98±1.06). Similarly, 76.5% stated that their anxiety/stress levels had increased due to new testing demands (M=3.86±1.01). Furthermore, 64.2% reported that their physical health had been affected. The Composite MPEI significantly exceeded the neutral threshold (M=3.35±0.812) (Table 3). A concerning finding was that 35.8% of staff reported considering leaving their position due to current job demands (2.59±1.10). Additional results from Table 4 indicate that 54.3% of staff agreed or strongly agreed that their mental health had been negatively impacted by the implementation of COVID-19 testing (M=3.37±1.13). Furthermore, nearly half of the respondents (46.9%) reported a decrease in overall job satisfaction due to the added responsibilities (M=2.99±1.16). A concerning finding was that 35.8% of staff reported considering leaving their position due to current job demands (2.59±1.10).

**Table 3:** Analysis of Mental and Physical Health Effects of Implementation (MPEI) post-COVID-19 responses of HIV Viral Load staff within public health sector laboratories in South Africa. Data was assessed using a likert scale (1 to 5), with the mean and standard deviation (SD) reported.

| Mental and Physical Health Effect on Implementation (MPEI) Question | Mean (SD) |
| --- | --- |
| My anxiety/stress levels have increased due to COVID-19 testing | 3.86 (1.01) |
| I have experienced burnout/fatigue due to COVID-19 testing | 3.98 (1.06) |
| My physical health has been affected due to COVID-19 testing | 3.28 (0.98) |
| My mental health has been affected due to COVID-19 testing | 3.37 (1.13) |
| My job satisfaction has decreased due to added responsibilities | 2.99 (1.16) |
| I have considered leaving my position due to the current job demands | 2.59 (1.10) |
| Composite Health Effects Score (MPEI) | 3.35 (0.812) |

COVID-19 Coronavirus Disease 2019 SD: Standard Deviation MPEI: Mental and Physical Health Effects of Implementation
Source: Author's own work

**Table 4:** Spearman’s Rho Correlation between post-Coronavirus Disease 2019 (COVID) occupational challenges assessed using Unified Crisis Strain (AIC) and adverse health outcomes assessed using Mental and Physical Health Effects of Implementation (MPEI). Responses were obtained from HIV viral load staff at public health sector laboratories in South Africa.

| Variables correlated | Correlation coefficient (Rho) | Significance (p) | Relationship strength |
| --- | --- | --- | --- |
| Post-COVID-19 challenges (AIC) and Mental and Physical health effect of implementation (MPEI) | 0.449 | <0.001 | Medium-to-Strong, Positive |

COVID-19 Coronavirus Disease 2019 Rho: Spearman's Correlation coefficient P: p-value AIC: Unified Crisis Strain MPEI: Mental and Physical Health Effects of Implementation

### Relationship between Challenges and Health Outcomes

Spearman’s rank-order correlation was conducted to assess the relationship between environmental challenges and staff wellbeing. The analysis yielded a highly statistically significant positive correlation between the overall AIC and the negative MPEI experienced by staff (rho = 0.449, *p* < 0.001). (Table 4).

## Discussion

The study confirmed that integrating high-volume COVID-19 testing protocols severely exacerbated pre-existing systemic weaknesses within South Africa’s public-sector VL laboratories. The demographic profile highlights that the burden fell on a highly skilled, critical cohort of experienced, middle-aged female medical technologists, a finding that is consistent with previous studies reporting a similar demographic distribution within this professional group.^[27]^ The severity of staff workload was the most critical finding, validating the concept of systemic fragility, where the workforce was operating near maximum capacity before the crisis.^[28]^ A local study reported that during waves one and two, COVID-19 testing represented up to 40% of VL volumes. A local study reported that during waves one and two, COVID-19 testing represented up to 40% of VL volumes.^[6]^

The convergence of pressures, including resource scarcity (PPE) ^[10]^ and infrastructure pressure fused into a single AIC factor. Interpreted through the HBM, these intensified challenges act as heightened perceived barriers, actively undermining the staff’s efforts to maintain safety and professional quality of life.

The high OECs translated directly into severe psychological distress and physical health impairment, confirmed by the Composite Health Effects Score. Beyond mental strain, most respondents reported that their physical health was negatively affected. These findings, characterised by striking rates of burnout/fatigue and anxiety/stress, align with the high levels of psychological and physiological consequences observed among global frontline health workers.^[15, 29]^

The statistically significant positive correlation is the most crucial finding, empirically confirming that operational strain is a direct and verifiable causative factor in staff distress and burnout. This evidence invalidates approaches focusing solely on individual resilience and mandates that policymakers address the structural causes of harm. The high ‘Perceived Severity’ of the occupational risk, combined with inadequate institutional support, diminished the staff’s ‘Perceived Benefits’ of remaining in the role. The resulting emotional exhaustion served as a profound, negative Cue to Action, leading directly to the critical negative behavioural outcome of withdrawal, manifested by experienced staff considering leaving their positions.

Elevated staff retention risks severely undermine South Africa’s national HIV/AIDS response and erode the resilience of its public health diagnostic systems against future emergencies.^[1, 30]^ The finding that operational strain results directly in psychological injury heightens the risk of diagnostic errors. ^[15]^ The findings necessitate policy actions that pivot from crisis management to strategic, pre-emptive investment in human capital.

To immediately address the confirmed structural causes of harm, a mandatory call for a paradigm shift is needed, requiring multiple recommendations. The first involves the development and funding of a formal Crisis Surge Staffing Model, including protocols for rapid hiring or the activation of a laboratory reservist corps to absorb sudden volume increases.^[31, 32]^ Laboratory services also need to formally institutionalise mental health support by implementing a continuous, confidential, and mandatory Employee Assistance Programme (EAP).^[33]^ Clear Fatigue Management Policies defining maximum shifts and mandatory rest periods must also be instituted. Services also need to maintain a secure, centralised buffer stock of critical PPE, reagents, and consumables specific to pandemic response, separate from routine HIV VL supplies. In addition, there is a need to clearly categorise COVID-19 and similar infectious diseases as high-risk occupational exposures^[34]^, with corresponding compensation and support mechanisms. Organisations need to institute regular, mandatory simulation-based training on safe specimen handling and appropriate use of PPE.

Beyond immediate policy changes, future research is essential to track the long-term consequences of the pandemic and enhance system resilience. Future research should specifically track this cohort using longitudinal studies to determine the actual attrition rate of experienced VL laboratory staff, which is crucial given the high intent-to-leave findings. ^[35]^ Comparative analyses between the public and private sectors and qualitative studies exploring managerial perspectives on resource allocation are also warranted.^[35]^ Comparative analyses between the public and private sectors and qualitative studies exploring managerial perspectives on resource allocation are also warranted. The study questionnaire could be amended to conduct an annual evaluation to compared to the baseline findings reported here. In addition, the questionnaire could be amended for other disciplines for broader use in other diagnostic settings.

### Limitations

The interpretation of the findings is constrained by three principal limitations: the cross-sectional design limits the ability to establish true causal relationships; reliance on self-reported data introduces the potential for common-method bias; and the sample drawn solely from the public health laboratory system restricts the generalisability of the results to other settings.

## Conclusions

This study confirmed the profound cost of systemic fragility on the VL diagnostic workforce. The core hypothesis that unmitigated systemic strain translates into significant psychological harm is confirmed by the highly significant correlation between operational challenges and negative health outcomes. Chronic systemic fragilities, primarily overwhelming workload and understaffing, acted as powerful, negative cues to action, driving psychological distress, burnout, and, critically, intentions to abandon the profession. The finding that around a third of experienced medical technologists considered leaving their posts represents a direct, catastrophic threat to the resilience of South Africa’s entire public health diagnostic system.

## Acknowledgements

This article is based on research originally conducted as part of Somayya Sarang’s Master of Public Health dissertation titled ‘Occupational and Environmental Challenges and Effects of COVID-19 Testing Implementation Experienced by HIV Viral Load Laboratory Staff within a Public Health Sector Laboratory in South Africa’, submitted to the Department of Environmental Health, University of Johannesburg in 2026. The dissertation is currently unpublished and not publicly available. The dissertation was supervised by Eunice Matingo-Mutava and Naseem Cassim. The dissertation was reworked, revised and adapted into a journal article for publication. The author confirms that the content has not been previously published or disseminated and complies with ethical standards for original publication. The researcher expresses heartfelt thanks to her supervisors, the VL staff of the NHLS, for their support and cooperation. Sincere thanks are also extended to the statistician, Teboho Moloi, for his expert guidance and instrumental support during the data analysis phase of this research.

## Author Contributions

SS (Developed and executed research, conducted data analysis, prepared the first draft and provided editorial input). EM & NC (supervision).

## Competing interests

The authors declare that they have no financial or personal relationship(s) that may have inappropriately influenced them in writing this article.

## Funding information

The authors received no financial support for the research, authorship, and/or publication of this article.

## Data availability

The authors do not have permission to share the study data.

## Disclaimer

The views expressed in this manuscript are those of the author(s) and not those of the University of the Witwatersrand, University of Johannesburg or the NHLS.

---

**Supplementary Table (S1):**
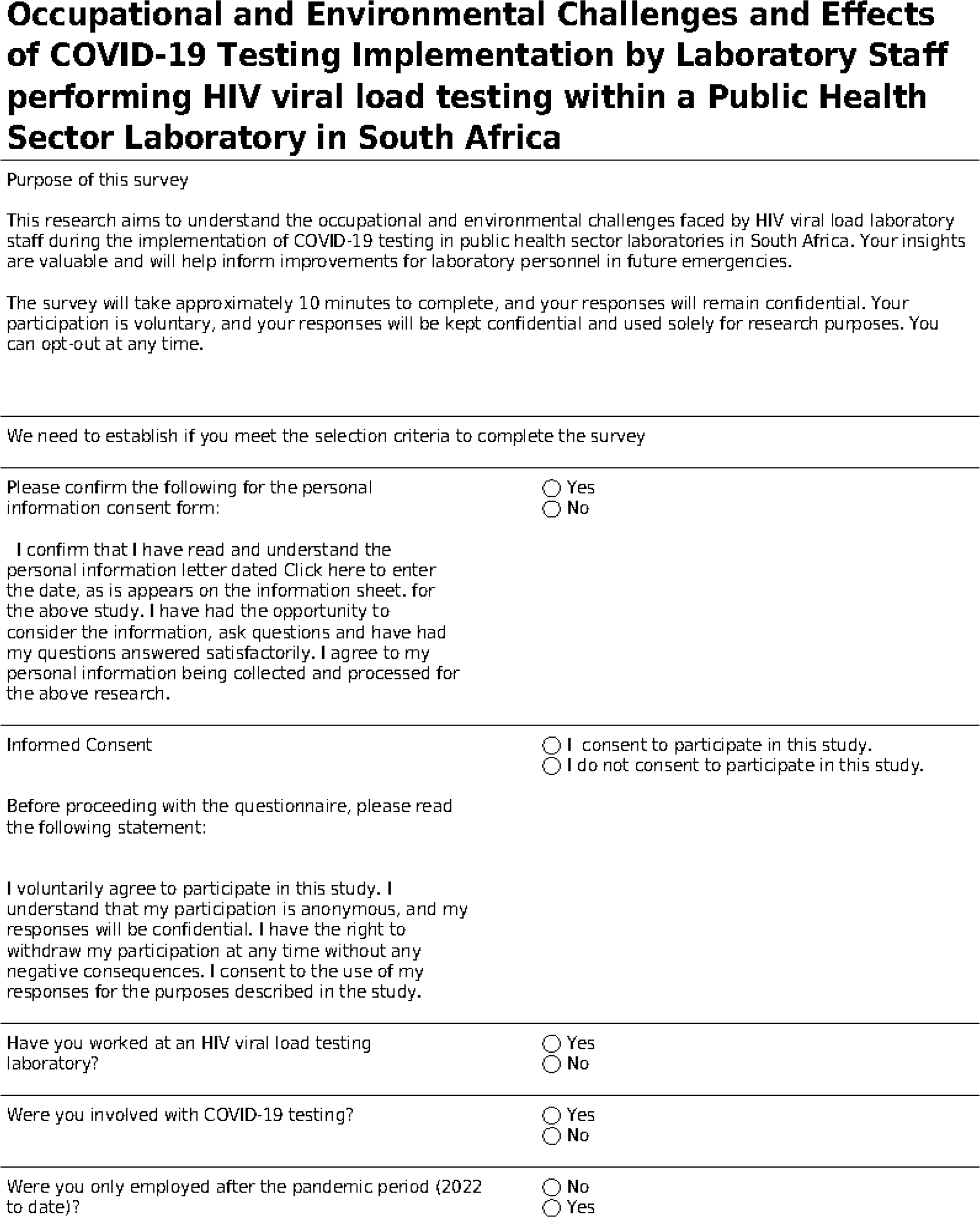
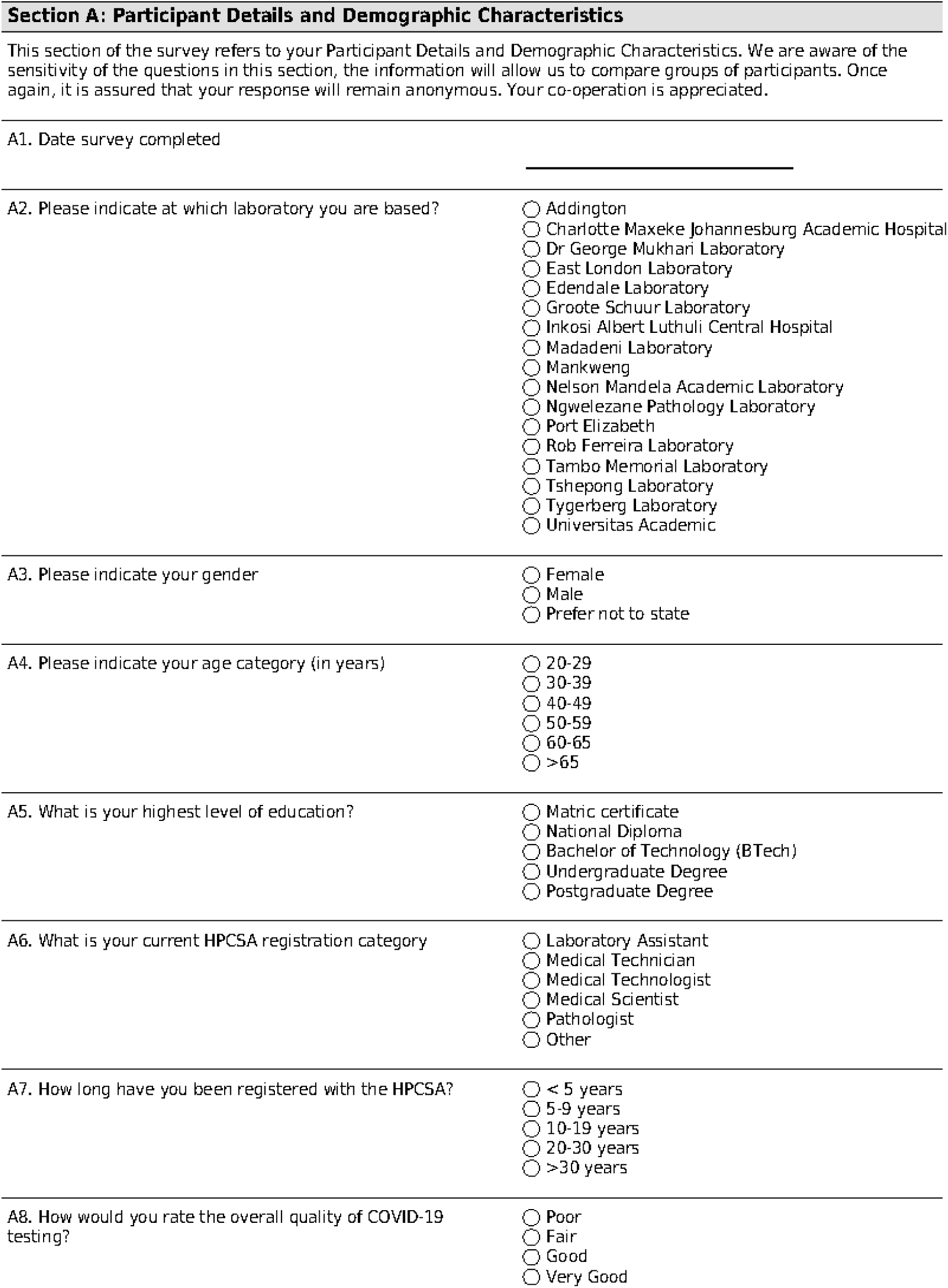
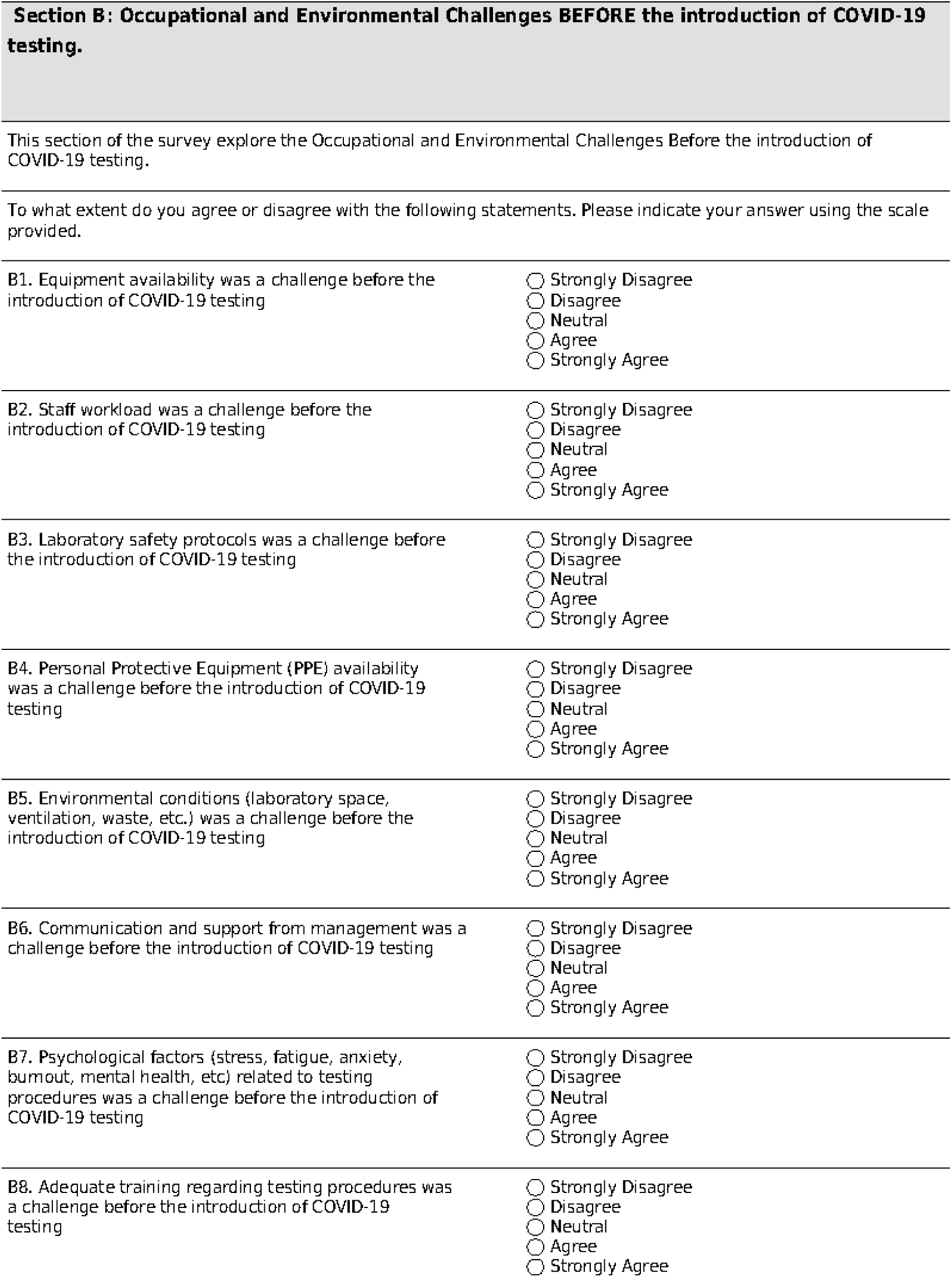
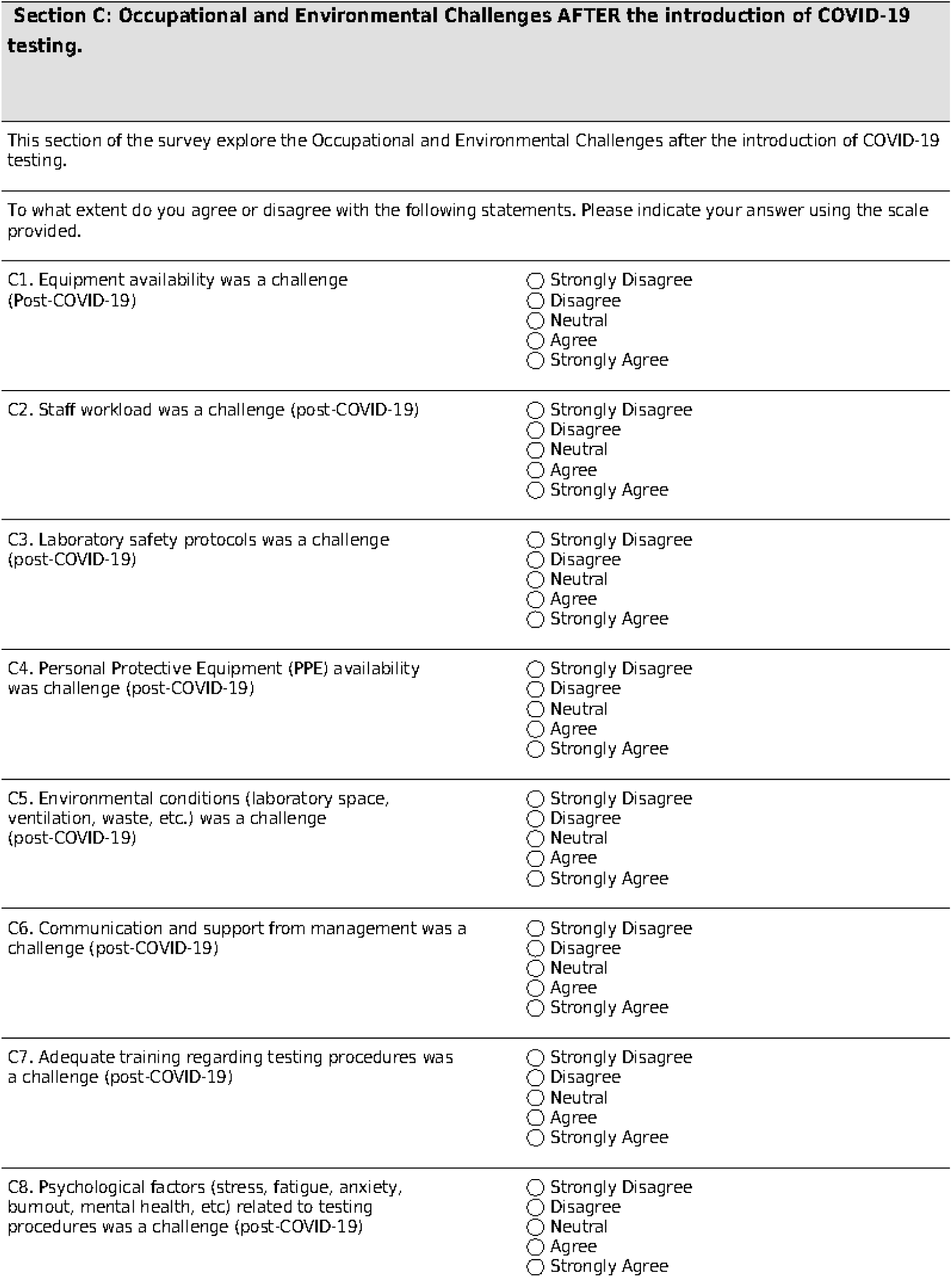
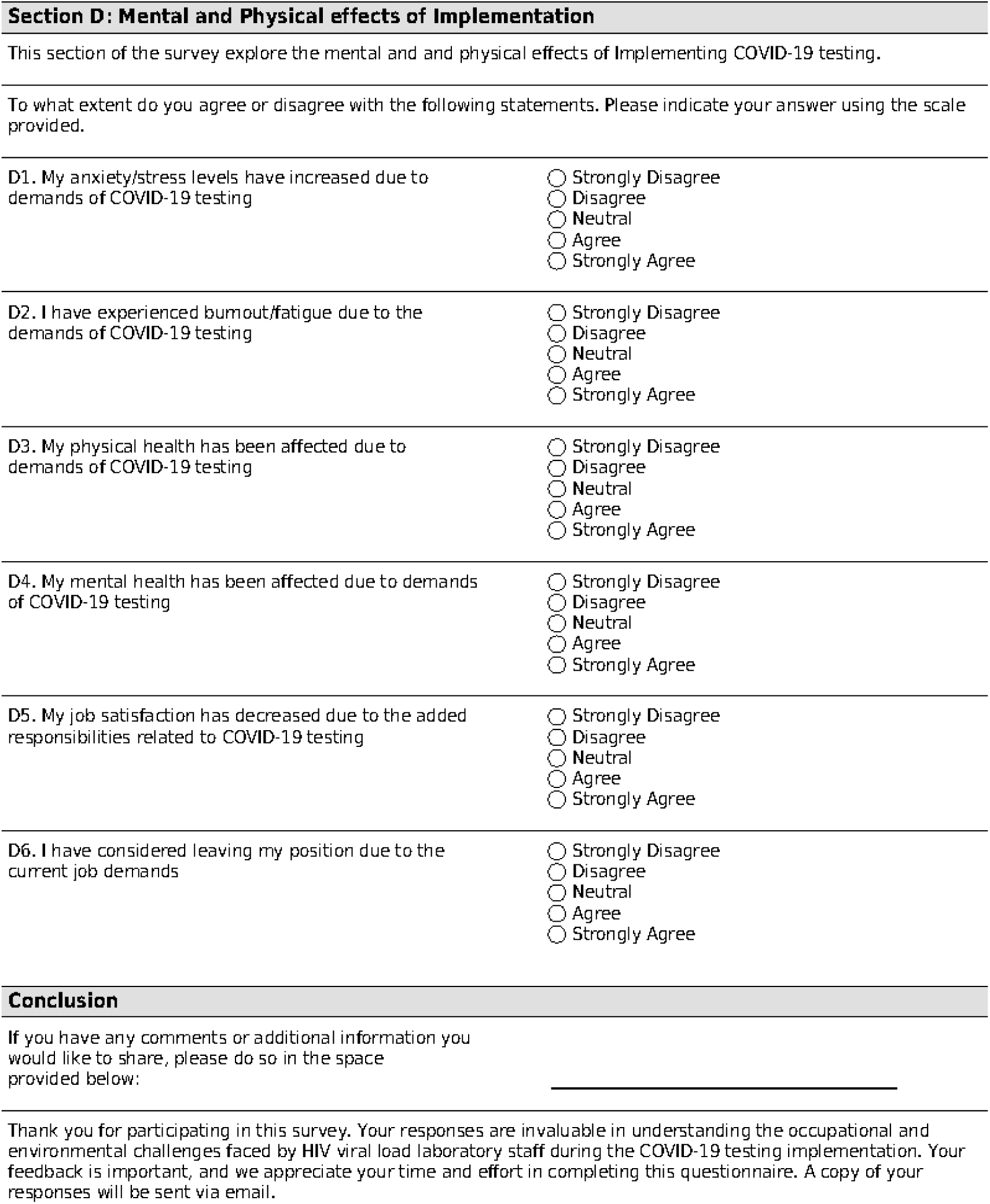
Self-administered REDCap electronic questionnaire.

## Notes

### Competing Interest Statement

The authors have declared no competing interest.

### Funding Statement

The author(s) received no specific funding for this work.

